# Evolution of antibodies against SARS-CoV-2 over seven months: experience of the Nationwide Seroprevalence ENE-COVID Study in Spain

**DOI:** 10.1101/2021.03.11.21253142

**Authors:** Mayte Pérez-Olmeda, José María Saugar, Aurora Fernández-García, Beatriz Pérez-Gómez, Marina Pollán, Ana Avellón, Roberto Pastor-Barriuso, Nerea Fernández-de Larrea, Mariano Martín, Israel Cruz, Jose L Sanmartín, Giovanni Fedele, Jose León Paniagua, Juan F Muñoz-Montalvo, Faustino Blanco, Raquel Yotti, Jesús Oteo-Iglesias, on behalf of the ENE-COVID Study Group

**Affiliations:** National Centre for Microbiology, Instituto de Salud Carlos III, Majadahonda, Madrid, Spain; Consortium for Biomedical Research in Epidemiology and Public Health (CIBERESP); National Centre for Epidemiology, Instituto de Salud Carlos III, Madrid, Spain; Deputy Directorate of Information Technologies, Ministry of Health, Madrid, Spain; National School of Public Health, Instituto de Salud Carlos III, Madrid, Spain; Instituto de Salud Carlos III, Madrid, Spain; General Secretary of Health, Ministry of Health, Madrid, Spain; Spanish Network for Research in Infectious Diseases (REIPI)

**Keywords:** COVID-19, antibodies, SARS-CoV-2, seroprevalence, ENE-COVID

## Abstract

**Objectives:** To analyse temporal trends in SARS-CoV-2 anti-nucleocapsid IgG throughout the four rounds of the nationwide seroepidemiologic study ENE-COVID (April-November 2020), and to compare the fourth-round results of two immunoassays detecting antibodies against nucleocapsid and to S protein receptor-binding domain (RBD).

**Methods:** A chemiluminescent microparticle immunoassay (CMIA) was offered to all participants in the first three rounds (Abbott; anti-nucleocapsid IgG). In the fourth round we offered this test and a chemiluminescence immunoassay (CLIA) (Beckman; anti-RBD IgG) to i) a randomly selected sub-cohort, ii) participants who were IgG-positive in any of the three first rounds; and iii) participants who were IgG-positive in the fourth round by point-of-care immunochromatography.

**Results:** Immunoassays involving 10,153 participants (82.2% of people invited to donate samples) were performed in the fourth round. A total of 2595 participants (35.1% of participants with immunoassay results in the four rounds) were positive for anti-nucleocapsid IgG in at least one round. Anti-nucleocapsid IgG became undetectable in 43.3% of participants with positive first-round results. Pneumonia was more frequent in participants with anti-nucleocapsid IgG in all four rounds (11.2%) than those in which IgG became undetectable (2.4%).

In fourth round, anti-nucleocapsid and anti-RBD IgG were detected in 5.5% and 5.4% participants of the randomly selected sub-cohort, and in 26.6% and 25.9% participants with at least one previous positive result, respectively. Agreement between techniques was 90.3% (kappa: 0.72).

**Conclusions:** The response of IgG to SARS-CoV-2 is heterogeneous and conditioned by infection severity. A substantial proportion of the SARS-CoV-2 infected population may have negative serologic results in the post-infection months.

## Introduction

As of February 14, 2021, SARS-CoV-2 had infected over 108 million people worldwide, causing over 2.3 million deaths [1]. Molecular testing based on specific nucleic acid amplification is the established method for early diagnosis of COVID-19 [2]. Most patients infected with SARS-CoV-2 develop antibodies to the surface spike (S) and nucleocapsid (N) proteins, which are therefore used as antigens in clinical serology assays. Such serologic assays are essential for developing and evaluating vaccines, antibody therapies, and serologic surveys [3]. However, current data regarding the longevity of antibodies to SARS-CoV-2 are inconsistent; some studies report a rapid decrease in specific IgG within approximately 3 months after infection [4,5], whereas others report IgG titers remaining stable over weeks or months [6-8].

Results from some serologic studies suggest differences in IgG behaviour depending on the virus protein to which it is directed; thus, some evidence [9,10] indicates that antibodies against N appear earlier than those directed against S but are less-protective against SARS-CoV-2 infection [10]. Titers of antibodies against SARS-CoV-2 appear to higher in patients with severe disease than in those with mild or asymptomatic disease [10,11], raising concerns about the impact of antibodies in the immune response to SARS-CoV-2.

Several SARS-CoV-2 serologic surveys have been conducted to estimate the proportion of the population exposed to SARS-CoV-2 and the durability of post-infection antibody production [6,12]. One such study is the ENE-COVID nationwide population-based longitudinal seroepidemiologic study in Spain [12]. Examining more than 60,000 randomly selected individuals over four rounds between April and November 2020, ENE-COVID covered the first and second pandemic waves in Spain. Serologic follow-up of a large cohort of participants was possible for 7 months. The general results revealed a national prevalence of 5–5.2% during the first wave of the pandemic (April–June 2020) [12,13], raising to 9.9% if we considering positive cases at any time between April and November [14].

The present study exploited the large and representative ENE-COVID project to i) analyse evolutionary trends in the detection of anti–N protein IgG using an immunoassay across the four rounds of the ENE-COVID study; and ii) describe the comparative serological results obtained in the fourth round using two different immunoassay formats to specifically detect anti–N protein and anti-RBD antibodies.

## Methods

### General study design and ENE-COVID study population

The ENE-COVID study is a nationwide, population-based cohort study of sero-prevalence, the general objectives of which were to i) estimate the prevalence of COVID-19 in the community-dwelling population of Spain by monitoring antibodies against SARS-CoV-2, and ii) evaluate evolutionary trends of antibodies over time. The design of ENE-COVID has been described elsewhere [12-14]. Briefly, 1,500 census tracts, with up to 24 households per tract, were randomly selected via two-stage sampling stratified by province and municipality size. The study invited around 95,000 people, including more than 68,000 participants in at least one of the first three rounds and around 51,000 in the last one.

The ENE-COVID study was developed in two phases during 2020; phase one included three rounds of analysis carried out during the first epidemic wave in Spain (April 27–May 11; May 18–June 1; June 8–June 22). Phase two included a fourth round developed during the second epidemic wave in the same cohort (November 16–29) (Figure 1).

**Figure 1.**
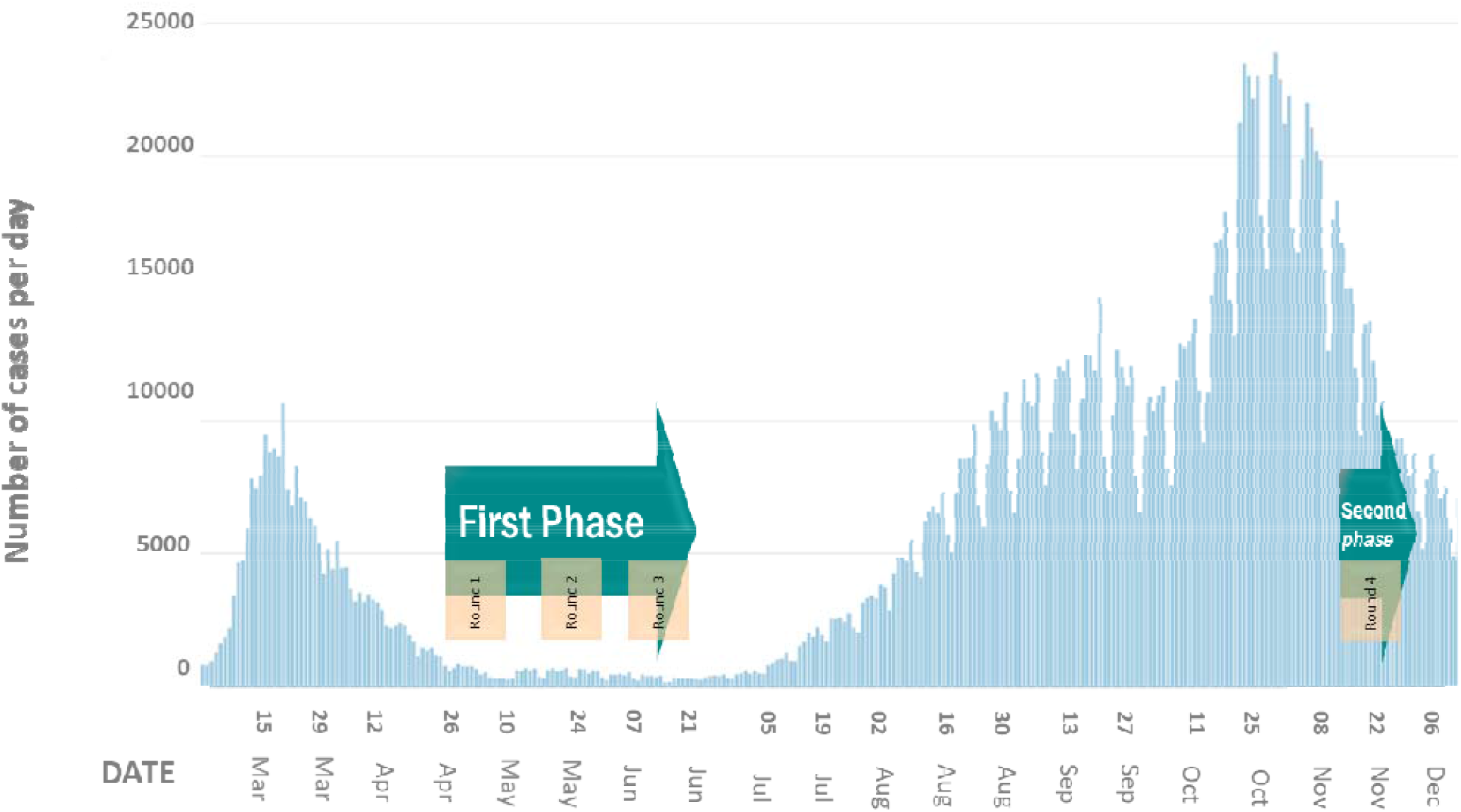
Epidemic curve of SARS-CoV-2 in Spain, with timeline of the four rounds of the ENE-COVID study. *Epidemic curve of the SARS-CoV pandemic: Data collated from individual data reported to the Red Nacional de Vigilancia Epidemiológica (RENAVE)*.

The Institutional Review Board of the Instituto de Salud Carlos III approved the study. Written informed consent was obtained from all participants.

### Serologic analyses

The serologic analyses carried out in ENE-COVID included direct rapid immunochromatography examinations of finger-prick blood samples to detect IgG/IgM against SARS-CoV-2 RBD (Orient Gene Biotech COVID-19 IgG/IgM, Orient Gene Biotech) in all participants, and two immunoassays that required venipuncture for subsequent laboratory analysis [12-14]. The immunoassays included a chemiluminescent microparticle immunoassay (CMIA) to detect anti–N protein IgG technique, and, in the fourth round, a chemiluminescence immunoassay (CLIA) to detect IgG against the RBD of S protein. The CMIA was used in all four rounds of the study, whereas the CLIA was used only in round four.

The SARS-CoV-2 IgG CMIA (Abbott Laboratories, Illinois, USA) allows qualitative detection of IgG directed against the nucleocapsid using serum obtained from venipuncture blood. Samples were tested on an ARCHITECT i2000SR high-performance analyser. According to the manufacturer’s data, the assay has 100% sensitivity and 99.6% specificity in confirmed cases 14 days after onset of symptoms. In a reliability study carried out at the National Centre of Microbiology (CNM), the CMIA exhibited 89.7% sensitivity and 100% specificity [12]. A meta-analysis of 23 studies evaluating this technique [15] reported a sensitivity of 90.6% and specificity of 99.3%.

The ACCESS SARS-CoV-2 CLIA (Beckman Coulter Inc., California, USA) allows the qualitative detection of IgG directed against S protein RBD using serum obtained from venipuncture blood. Samples were tested on a UniCel Dxl 800 high-performance analyser. The assay’s sensitivity and specificity as reported by the manufacturer in confirmed cases 14 days after onset of symptoms are 99.1% and 99.8%, respectively. In a reliability study carried out at the CNM, the CLIA exhibited a sensitivity of 98.8% and specificity of 100% (Supplementary Table S1). Other studies have reported a sensitivity of approximately 82% in confirmed cases >14 days after onset of symptoms [16,17].

The present study reports immunoassay serology results obtained using both the Abbott and Beckman assays.

### Selection of participants for immunoassay analyses

Samples from all participants in the ENE-COVID study who agreed to donate a blood sample (>85%) were examined using the Abbott CMIA in the first three rounds. In the fourth round, both immunoassays (Abbott CMIA of and Beckman CLIA) were used for serologic analyses of patient samples. However, blood sample collection in the fourth round was limited to certain sub-groups of participants, as follows: a) a randomly selected sub-cohort of 15% of the ENE-COVID cohort; b) participants who had an IgG-positive result in any of the three first rounds either by CMIA or using the above-mentioned rapid immunochromatography test; and c) participants who had a fourth-round IgG-positive result by the rapid immunochromatography test [14]. Data are included in this report for all participants who had CMIA results in the fourth round of the ENE-COVID study.

### Statistical analyses

The percentage of positive results by rounds, with 95% confidence intervals (CI), was calculated. The level of agreement between the tests was evaluated using Cohen’s kappa score [18]. Statistical analyses were performed using GraphPad Prism software v.7.02 (GraphPad Software Inc., San Diego, CA, USA).

## Results

### Evolution of results for IgG against N (Abbott CMIA) across the four rounds of ENE-COVID

In the fourth round of the ENE-COVID study, blood samples were drawn by venipuncture from a total of 10153 participants (82.2% of the participants invited to donate a blood sample).

Abbott CMIA results were available for all four rounds in 7400 (72.9% of those with CMIA in the fourth round) participants. Of these participants, 2595 (35.1%) had a positive result in at least one of the four rounds. Of this sub-group, 537 (20.7%) maintained detectable IgG levels across all four rounds, 875 (33.7%) did not have an IgG-positive result in the first round but did exhibit positive results in later rounds, and 887 (34.2%) had detectable IgG in the first round, but the levels declined to undetectable during the study (Table 1). The remaining 11.4% of this sub-group presented atypical result sequences over the four rounds of ENE-COVID, with negative/negative/positive/negative (n=163; 6.3% of all cases with at least one positive result) and positive/positive/negative/positive (n=93; 3.6% of all cases with at least one positive result) results sequences predominating.

**Table 1.** Evolution of IgG against SARS-CoV-2 nucleocapsid (N) protein in the four rounds of the ENE-COVID study (only participants with immunoassays results in the four rounds are included)

| **Participants** | **Number (%; CI 95%)** |
| --- | --- |
| *At least one positive IgG determination in any of the rounds* | 2595 |
| Positive result in all four rounds | 537 (20.7; 19.1-22.3)* |
| Evolution to seropositive anti-N IgG | 875 (33.7; 31.9-35.6)* |
| Evolution to seronegative anti-N IgG | 887 (34.2; 32.3-36.0)* |
| Atypical antibody evolution** | 256 (9.9; 8.7-11.1)* |
| *Positive result for anti-N IgG in the first round* | 1530 |
| Evolution to seronegative anti-N IgG | 887 (58; 55.5-60.5)*** |
| Evolution to seronegative anti-N and anti-RBD IgG | 662 (43.3; 40.8-45.8)*** |

\*Percentages referred to the total number of cases with at least one positive result in any of the four rounds.
\*\*Includes results with atypical evolution (see text).
\*\*\*Percentages referred to the total number of cases with positive IgG result in the first round

Fifty-eight percent of participants (887/1530) who had a positive IgG result for N protein in the first round evolved to seronegative for these antibodies throughout the study (Table 1). Of these participants, 25.4% had a positive Beckman CLIA result for IgG against the S protein RBD in the fourth round. Excluding these cases, in 43.3% of participants positive for IgG to the N protein in the first round, neither IgG for N (Abbott CMIA) nor IgG for the RBD (Beckman CLIA) were detected in the fourth round (sero-reversion) (Table 1). As expected, the highest number of sero-reversions occurred between the third and fourth rounds (467 cases, representing 70.5% of all sero-reversion cases).

The percentage of participants who developed pneumonia was higher in patients who were positive for IgG against N across all four rounds (11.2% [60/537]) than in patients in which IgG against both N and the RBD of S became undetectable during the study (2.4% [16/662]). Among participants with atypical result sequences, 11.8% of those with positive/positive/negative/positive results developed pneumonia, where only 1.2% of patients with negative/negative/positive/negative results developed pneumonia.

### Results of the fourth round of ENE-COVID

In the fourth round of the ENE-COVID study, serum samples of 10153 participants were analysed using two high-performance serologic techniques. A total of 2032 participants met more than one inclusion criteria.

Table 2 summarizes the results of the Abbott CMIA (IgG against N protein) and Beckman CLIA (IgG against the RBD of the S protein) in the participants of the fourth round of the ENE-COVID study, classified according to the different sub-groups that were invited to blood collection.

**Table 2.** Comparison of results of the Abbott (anti–N protein) and Beckman (anti-RBD) immunoassays performed in the fourth round of the ENE-COVID study.

|  | **All participants,**<br>**number (%; CI 95%)** | **Participants of the**<br>**randomly selected sub-**<br>**cohort, number (%; CI**<br>**95%)** | **Participants with at least**<br>**one positive result in the**<br>**first three rounds, number**<br>**(%; CI 95%)** | **Participants who had a**<br>**positive result by the rapid**<br>**test in the fourth round,**<br>**number (%; CI 95%)** |
| --- | --- | --- | --- | --- |
| **Total\*** | 10153 | 5827 | 3261 | 3263 |
| **Anti-N IgG positive** | 2220 (21.9; 21.1-22.7) | 321 (5.5; 4.9-6.1) | 867 (26.6; 25.1-28.1) | 1903 (58.3; 56.7-60.0) |
| **Anti-RBD IgG**<br>**positive** | 2191 (21.6; 20.8-22.4) | 315 (5.4; 4.8-6.0) | 846 (25.9; 24.4-27.4) | 2040 (62.5; 60.9-64.2) |
| **Anti-N and -RBD**<br>**IgG positive** | 1713 (16.9; 16.1-17.6) | 248 (4.3; 3.7-4.8) | 467 (14.3; 13.1-15.5) | 1648 (50.5; 48.8-52.2) |
| **Anti-N IgG**<br>**positive/Anti-RBD**<br>**IgG negative** | 507 (5.0; 4.6-5.4) | 73 (1.2; 1.0-1.5) | 400 (12.3; 11.1-13.4) | 255 (7.8; 6.9-8.7) |
| **Anti-N IgG**<br>**negative/Anti-RBD**<br>**IgG positive** | 478 (4.7; 4.3-5.1) | 67 (1.1; 0.9-1.4) | 379 (11.6; 10.5-12.7) | 391 (12.0; 10.9-13.1) |
| **Anti-N and -RBD**<br>**IgG negative** | 7455 (73.4; 72.6-74.3) | 5439 (93.3; 92.7-94.0) | 2415 (74.1; 72.6-75.6) | 969 (29.7; 28.1-31.3) |
| **Agreement (%)** | 90.3 | 97.6 | 88.4 | 88.2 |

*N: nucleocapsid; RBD: receptor-binding domain. \* There are 2032 participants included in more than one group.*

In the participants included in the randomly selected sub-cohort (n=5827), positive IgG results were obtained for 321 (4.9%) and 315 (5.4%) participants by the Abbott and Beckman immunoassays, respectively. Among participants with at least one positive result in any of the three first rounds (n=3261), 867 (26.6%) and 846 (25.9%) participants had a positive result for IgG against N (Abbott CMIA) and the RBD of S (Beckman CLIA), respectively. These figures were 1093 (58.3%) and 2040 (62.5%) by Abbott and Beckman immunoassays, respectively, in the sub-cohort of participants who had a positive result by the rapid test in the fourth round (n=3263).

These high-performance immunoassays exhibited 90.3% agreement, with a Kappa index of 0.72 (95% CI: 0.70–0.73). Cases in which there was lack of agreement between the CMIA and CLIA (n = 985; 9.7%) were distributed almost equally between those with a positive result for IgG against N (Abbott CMIA) and negative result for IgG against the RBD of S (Beckman CLIA) (51.5%), and vice versa (48.5%). In the fourth round, agreement between rapid test and CMIA was 83.5% (Kappa index: 0.58; 95% CI: 0.56-0.60), and between rapid test and CLIA was 86.4% (Kappa index: 0.66; 95% CI: 0.64-0.67).

Participants who had positive results by both immunoassays in the fourth round suffered pneumonia more frequently (11.3% [194/1713]) than participants who had only one positive immunoassay result in the fourth round (5.8% [57/985]).

## Discussion

Two important findings emerged from the results of the present study. First, our data suggest that a substantial percentage of the population infected with SARS-CoV-2 may exhibit negative serologic test results in the months following infection. Second, we observed heterogeneity in the immunologic response regarding production of IgG against either the SARS-CoV-2 N protein or S protein RBD. These data were derived from analyses of a large cohort of non-hospitalized participants randomly selected from the general population tested four times over a period of 7 months.

Declines in the levels of antibodies to SARS-CoV-2 in the months following infection have been described by previous studies involving smaller populations [11,19]. In a recent study of 156 healthcare personnel in the USA [19], 93.6% exhibited a decrease in antibody levels after 60 days, and in 28.2% of cases, IgG against SARS-CoV-2 became undetectable. Sero-reversion occurred in 50% of asymptomatic infected individuals in that study [19]. Our representative population study shows an evolution toward un-detectability of IgG over the 7 months of the study in 43.3% of participants with positive first-round results. However, this finding is not necessarily indicative of a reduction in immunity against SARS-CoV-2. Although protective immunity against SARS-CoV-2 is not well defined in humans, the immune memory associated with memory T and B cells could generate long-term protective immunity, as occurs with other infectious diseases [20,21]. Another study that examined different indicators of circulating immune memory to SARS-CoV-2 in 188 COVID-19 patients [11] detected at least three indicators of immunologic memory in 95% of participants with 5–8 months of symptom onset, indicating that long-lasting immunity against a second SARS-CoV-2 infection is a real possibility in most individuals. Indeed, although cases of re-infection have been documented [22,23], they are rare from a global epidemiologic perspective. Studies carried out specifically to identify cases of symptomatic re-infection in large cohorts of patients did not report any such cases [24,25].

The lower frequency of pneumonia among those in which IgG levels became undetectable in the present study is consistent with observations confirmed in recent studies [11,15,26,27].

Recent studies described the predominance of S-specific versus N-specific antibodies in individuals with mild versus severe disease, respectively [10,28]. This difference suggests that a strong humoral response to S could limit the effect of viral infection. In the ENE-COVID study, no association between increased disease severity and an imbalance in humoral immunity to the N protein versus the RBD of the S protein was observed.

Although antibodies against N appear earlier than antibodies against S [9,10], the latter seem to be more stable over time. Bearing this in mind, the discordance between the detection of IgG against N versus IgG against the RBD of S may be associated with how recent infection occurred, such that IgG against the RBD of S were not yet detectable in cases of more recent infection, or in cases of long evolution after infection, in which levels of IgG against the N protein had decreased to un-detectability. Alternatively, these apparent discrepancies could also be explained by the heterogeneous antibody response of COVID-19 patients, likely involving various as yet unidentified factors, in addition to disease severity [11,27]. In our study, development of pneumonia was correlated with the simultaneous presence of IgG against both N and the RBD of S.

A number of cases in the present study (n=256; 9.9%) with atypical result sequences across the four rounds were mainly due to discrepant results in the third round with respect to the other three rounds. Taking into account the temporal distribution of the ENE-COVID rounds in relation to the first waves of the pandemic in Spain (Figure 1), these cases could be explained by several scenarios: i) antibody levels at the detection limit thresholds of the serologic assays used in the study, ii) mild infections in the third round in which the level of antibodies decreased in the fourth round, or iii) cases with a new contact with the virus between the third and fourth rounds, which would have led to reactivation of the immune system via memory cells. It should be noted that the high percentage of patients developing reporting pneumonia (11.8%) among cases of positive determinations in all rounds except the third round was very similar to that of cases with positive determinations in all rounds (11.2%).

Our data show two remarkable findings: i) a substantial percentage of SARS-CoV-2–infected patients may have negative serologic test results in the months following infection, and ii) the serologic IgG response to SARS-CoV-2 targets is heterogeneous and conditioned by disease severity.

## Data Availability

N/A

## Transparency Declaration

The authors have none to declare.

## Contribution of authors

RY, FB, JFM-M, MP and JO-I conceived and designed the study. MM, JLS, RY, FB, MP, IC, JLP and JO-I coordinated the study. IC and JLP gave training and logistical support to the study. MP-O, JMS, AF-G, AA and ENE-COVID Study Group performed the experiments. MM, JLS, BP-G, MP, NF-D, RP-B, AA and MP-O created the databases and analyzed the results. MP-O, AA, AF-G and GF created de serum biobank. MP-O and JO-I wrote the manuscript. All authors have read, edited and approved the final manuscript.

## Acknowledgments

Members of the ENE-COVID Study Group are listed in Supplementary material.

