## Supplemental Table 1 for "Evolution of antibodies against SARS-CoV-2 over seven months: experience of the Nationwide Seroprevalence ENE-COVID Study in Spain"

**Contents	Page**

ENE-COVID Study Group	2

Supplementary Table S1	6

**ENE-COVID STUDY GROUP***

* Names listed alphabetically within each institution or regional health service.

**Spanish Ministry of Health**

Pilar Aparicio Azcárraga; Faustino Blanco; Rodrigo Gutiérrez Fernández; Mariano Martín; Saturnino Mezcua Navarro; Marta Molina; Juan F. Muñoz-Montalvo; Matías Salinero Hernández; Jose L. Sanmartín.

**Instituto de Salud Carlos III**

Manuel Cuenca-Estrella; José León Paniagua; Raquel Yotti.

National Center of Epidemiology: Nerea Fernández de Larrea, Centro de Investigación Biomédica en Red de Epidemiología y Salud Pública (CIBERESP); Pablo Fernández-Navarro, CIBERESP; Olivier Nuñez, CIBERESP; Roberto Pastor-Barriuso, CIBERESP; Beatriz Pérez-Gómez, CIBERESP; Marina Pollán, CIBERESP.

National Center of Microbiology: Ana Avellón, CIBERESP; Beatriz Bellido; Giovanni Fedele; Aurora Fernández-García, CIBERESP; Jesús Oteo-Iglesias, Spanish Network for Research in Infectious Diseases (REIPI); María Teresa Pérez Olmeda; José María Saugar.

National School of Public Health: Israel Cruz; Maria Elena Fernández Martínez; Francisco D. Rodríguez-Cabrera.

**Spanish Regional Health Services**

***Andalucía***

Health Services: Susana Padrones Fernández, Distrito Sanitario Sevilla; José Manuel Rumbao Aguirre, Distrito Sanitario Córdoba Guadalquivir.

Laboratory: José M. Navarro Marí, Hospital Universitario Virgen de las Nieves, Instituto Biosanitario ibs.Granada; Begoña Palop Borrás, Hospital HRU de Málaga; Ana Belén Pérez Jiménez, Hospital Universitario Reina Sofía, Instituto Maimónides de Investigación Biomédica de Córdoba (IMIBIC); Manuel Rodríguez-Iglesias, Hospital Universitario Puerta del Mar-INIBICA, Cádiz.

***Aragón***

Health Services: Ana María Calvo Gascón, Servicio Aragonés de Salud; María Luz Lou Alcaine, Gobierno de Aragón.

***Asturias***

Health Services: Ignacio Donate Suárez, Consejería de Salud; Oscar Suárez Álvarez, Servicio de Salud del Principado de Asturias.

Laboratory: Mercedes Rodríguez Pérez, Hospital Universitario Central de Asturias (HUCA).

***Baleares***

Health Services: Margarita Cases Sanchís, Servei de Salut de les Illes Balears; Carlos Javier Villafáfila Gomila, Servei de Salut de les Illes Balears.

Laboratory: Lluis Carbo Saladrigas, Hospital Mateu Orfila; Adoración Hurtado Fernández, Hospital Can Misses; Antonio Oliver, Hospital Universitario Son Espases, Instituto de Investigación Sanitaria Illes Balears (IdISBa), REIPI.

***Canarias***

Health Services: Elías Castro Feliciano, GSA 112; María Noemí González Quintana, Gestión de Servicios para la Salud y Seguridad en Canarias (GSC).

Laboratory: José María Barrasa Fernández, Hospital Universitario Nuestra Señora de la Candelaria; María Araceli Hernández Betancor, Hospital Universitario Insular de Gran Canaria; Melisa Hernández Febles, Hospital General de Gran Canaria Dr. Negrín; Leopoldo Martín Martín, Hospital de La Palma.

***Cantabria***

Health Services: Luis-Mariano López López, Servicio Cántabro de Salud; Teresa Ugarte Miota, Servicio Cántabro de Salud.

Laboratory: Inés De Benito Población, Hospital Sierrallana.

***Castilla-La Mancha***

Health Services: María Sagrario Celada Pérez, Servicio de Salud de Castilla-La Mancha; María Natalia Vallés Fernández, Servicio de Salud de Castilla-La Mancha.

***Castilla y León***

Health Services: Tomás Maté Enríquez, Gerencia Regional de Salud de Castilla y León; Miguel Villa Arranz, Gerencia Regional de Salud de Castilla y León.

Laboratory: Marta Domínguez-Gil González, Hospital Universitario Río Hortega; Isabel Fernández-Natal, Complejo Asistencial Universitario de León; Gregoria Megías Lobón, Complejo Asistencial Universitario de Burgos; Juan Luis Muñoz Bellido, Complejo Asistencial Universitario de Salamanca.

***Cataluña***

Health Services: Pilar Ciruela, Departament de Salut, CIBERESP; Ariadna Mas i Casals, Departament de Salut.

Laboratory: Maria Doladé Botías, Laboratorio Clínic Metropolitana Nord, Hospital Universitari Germans Trias i Pujol; M. Angeles Marcos Maeso, Centro de Diagnóstico Biomédico, Hospital Clínic de Barcelona, REIPI; Dúnia Pérez del Campo, Laboratorio Territorial de Girona.

***Comunidad Valenciana***

Health Services: Antonio Félix de Castro, Conselleria de Sanitat Universal i Salut Pública; Ramón Limón Ramírez, Conselleria de Sanitat Universal i Salut Pública.

***Extremadura***

Health Services: Maria Francisca Elías Retamosa, Servicio Extremeño de Salud; Manuela Rubio González, Servicio Extremeño de Salud.

***Galicia***

Health Services: María Sinda Blanco Lobeiras, Sergas; Alberto Fuentes Losada, Consellería de Sanidade.

Laboratory: Antonio Aguilera, Complexo Hospitalario Universitario de Santiago de Compostela (CHUS); German Bou, Hospital Universitario A Coruña, REIPI.

***La Rioja***

Health Services: Yolanda Caro, Hospital San Pedro; Noemí Marauri, Atención Primaria.

Laboratory: Luis Miguel Soria Blanco, Hospital San Pedro.

***Madrid***

Health Services: Isabel del Cura González, Gerencia Asistencial Atención Primaria, Universidad Rey Juan Carlos, Red de Investigación Servicios de Salud en Enfermedades Crónicas (REDISSEC); Montserrat Hernández Pascual, Gerencia Adjunta Procesos Asistenciales Atención Primaria.

Laboratory: Roberto Alonso Fernández, Hospital General Universitario Gregorio Marañón; Paloma Merino-Amador, Hospital Universitario Clínico San Carlos.

***Murcia***

Health Services: Natalia Cabrera Castro, Consejería de Salud, IMIB-HCUV Arrixaca, Servicio Murciano de Salud; Aurora Tomás Lizcano, Servicio Murciano de Salud.

Laboratory: Cristóbal Ramírez Almagro, Hospital General Universitario Santa Lucía; Manuel Segovia Hernández, Hospital Clínico Universitario Virgen de la Arrixaca, Universidad de Murcia.

***Navarra***

Health Services: Nieves Ascunce Elizaga, Public Health and Labour Institute of Navarre, CIBERESP; María Ederra Sanz, Public Health and Labour Institute of Navarre, CIBERESP.

Laboratory: Carmen Ezpeleta Baquedano, Complejo Hospitalario de Navarra.

***País Vasco***

Health Services: Ana Bustinduy Bascaran, Dirección General de Osakidetza; Susana Iglesias Tamayo, Dirección General de Osakidetza.

Laboratory: Luis Elorduy Otazua, Hospital Universitario Cruces.

***Ceuta***

Health Services: Rebeca Benarroch Benarroch, Consejería de Sanidad, Consumo y Gobernación; Jesús Lopera Flores, Instituto Nacional de Gestión Sanitaria.

***Melilla***

Health Services: Antonia Vázquez de la Villa, Instituto Nacional de Gestión Sanitaria.

**Supplementary Table S1.** Specificity and sensitivity of the Beckman Coulter CLIA (detection of IgG against the S protein RBD) determined at the Serology Laboratory of the National Centre Microbiology.

| **ACCESS SARS-CoV-2 IgG** | **PCR-positive samples ≥10 days after onset of symptoms** | **Negative controls (Specificity)**** |
| --- | --- | --- |
| **Positive** | 80* | 0 |
| **Negative** | 1 | 38 |
| **Total** | 81 | 38 |
| Sensitivity: 98.8% (95% CI: 93.3–100) | | |
| Specificity: 100% (95% CI: 90.75–100) | | |

**Two samples with equivocal results in the commercial assays were regarded as reactive (positive) in this analysis.*

***Negative serum sample panel consisting of samples collected retrospectively prior the SARS-CoV-2 epidemic (November 2019).*
